# Global associations of maternal hypertensive disorders and offspring allergic disease burden

**DOI:** 10.1101/2024.04.29.24306588

**Authors:** Duan Ni, Ralph Nanan

## Abstract

**Objectives:** Maternal hypertensive disorders (MHD) are widespread globally, modifying maternal and fetal immunity, and have been linked to increased allergic diseases in offsprings. Nevertheless, so far, most studies in this field are small-scale and results remain inconclusive.

**Methods:** Harnessing unprecedented global allergic disease and pregnancy data covering more than 150 countries from 1990 to 2019 as proxies, we leveraged the state-of-the-art generalized additive model (GAM) to interrogate the potential link between MHD and common offspring allergic diseases, exemplified by atopic dermatitis (AD) and asthma.

**Results:** A model considering the main effects from MHD, socioeconomic factor like GDP and time, as well as their interactions was favoured, suggesting their interactive effects on offspring allergic diseases. Generally, MHD in pregnancies were associated with increased AD and asthma in offsprings early in life, and a more pronounced effect was found for AD relative to asthma.

**Conclusions:** Globally, MHD in pregnancies are linked to increased offspring allergic disease burden, which, with further in-depth investigations, would inform allergic disease preventions in clinic. Our analyses also support the Developmental Origins of Health and Disease (DOHAD) concept and showcase a novel methodology for DOHAD-related research.

---

Maternal hypertensive disorders (MHD) are widespread globally, exhibiting a spectrum of severity from pregnancy-induced hypertension to preeclampsia. MHD alter maternal and fetal immunity and have been linked to increased allergic diseases in offspring, although most studies are small-scale, with inconclusive results^1,2^. Harnessing comprehensive global data, we systematically assessed the correlations between MHD and common offspring allergic diseases, exemplified by atopic dermatitis (AD) and asthma.

We focused on 1–4-year-olds, the peak incidence ages for AD and asthma^3^. Ratios of 1-4 years old AD or asthma cases versus number of pregnancies 2 years prior for each country were calculated. These ratios were used as proxies of percentages of pregnancies with offsprings developing AD (AD%) or asthma (asthma%) at 2 years of age. These results were then compared with percentages of MHD-affected pregnancies (MHD%) at 2 years prior timepoint (Figure 1A). They were modelled as close proxies of the links between MHD and offspring allergic diseases. Additionally, socioeconomic status, reflected by gross domestic product (GDP), was accounted as a potential confounder^3^.

**Figure 1.**
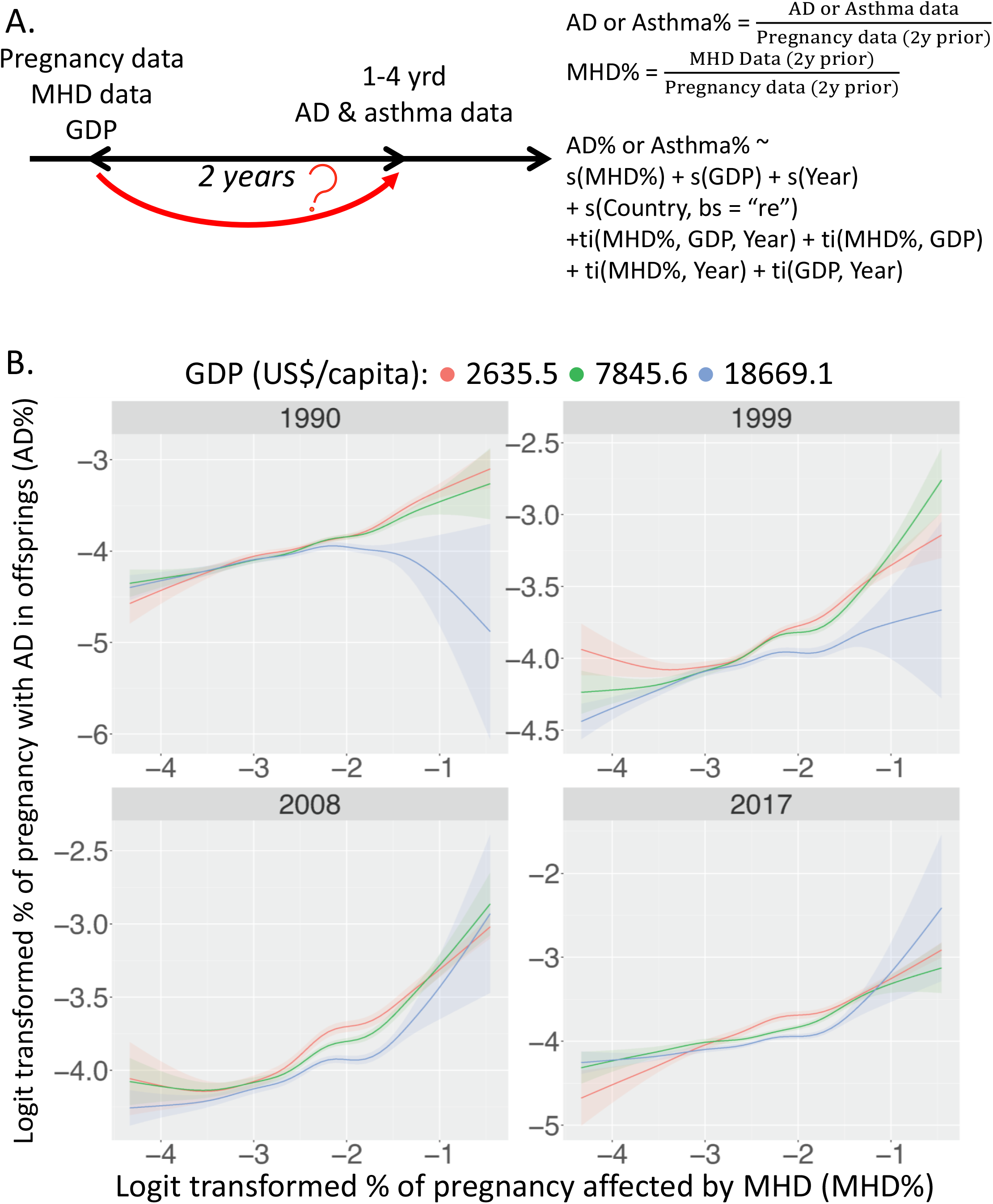
**A**. Percentages of pregnancies with offspring developing atopic dermatitis (AD%) or asthma (asthma%) at 2 years of age were calculated as the ratios of 1-4 years old AD or asthma cases versus pregnancies 2 years prior of the same country. These data were modelled with percentages of maternal hypertensive disorders-affected pregnancies (MHD%) 2 years prior, adjusting for gross domestic product (GDP) and time, as proxies of the link between MHD and offspring allergic diseases. A model considering the main effects from MHD%, GDP and time with their interactive effects were favoured. **B**. Predicted effects of logit transformed MHD%, GDP and time on the logit transformed AD%. (See Supplementary Information for statistics)

We leveraged the state-of-the-art generalized additive model (GAM) for analysis^4^, given its power to comprehensively survey multiple parameters, their interactions and non-linear effects. A series of GAMs were modelled for asthma% or AD%, using MHD%, GDP and time as predictors. Countries from which the data originated were adjusted as random effects (Supplementary Information). A model combining the main effects from MHD%, GDP and time, and their interactions was favoured (Supplementary Information), suggesting their interactive effects on offspring allergic diseases.

Figure 1B illustrates the predicted response curves of AD% as a function of MHD%. Four plots represent different time points (1990, 1999, 2008, 2017) and are stratified by GDP quantiles (red, 25%; green, 50%; blue, 75%). Generally, MHD% were associated with elevated AD%, except in 1990 within a high GDP setting. Otherwise, AD% minimally fluctuated with time and GDP.

MHD% were also generally associated with increased asthma%, albeit weaker than AD% (Figure 2). Notably, in 1990, 2008, and 1999, asthma% dropped as logit MHD% increased beyond −1 (MHD%∼26.9%), coupled with low GDP, suggesting their interactive effects. Otherwise, GDP and time mildly impacted asthma%.

**Figure 2.**
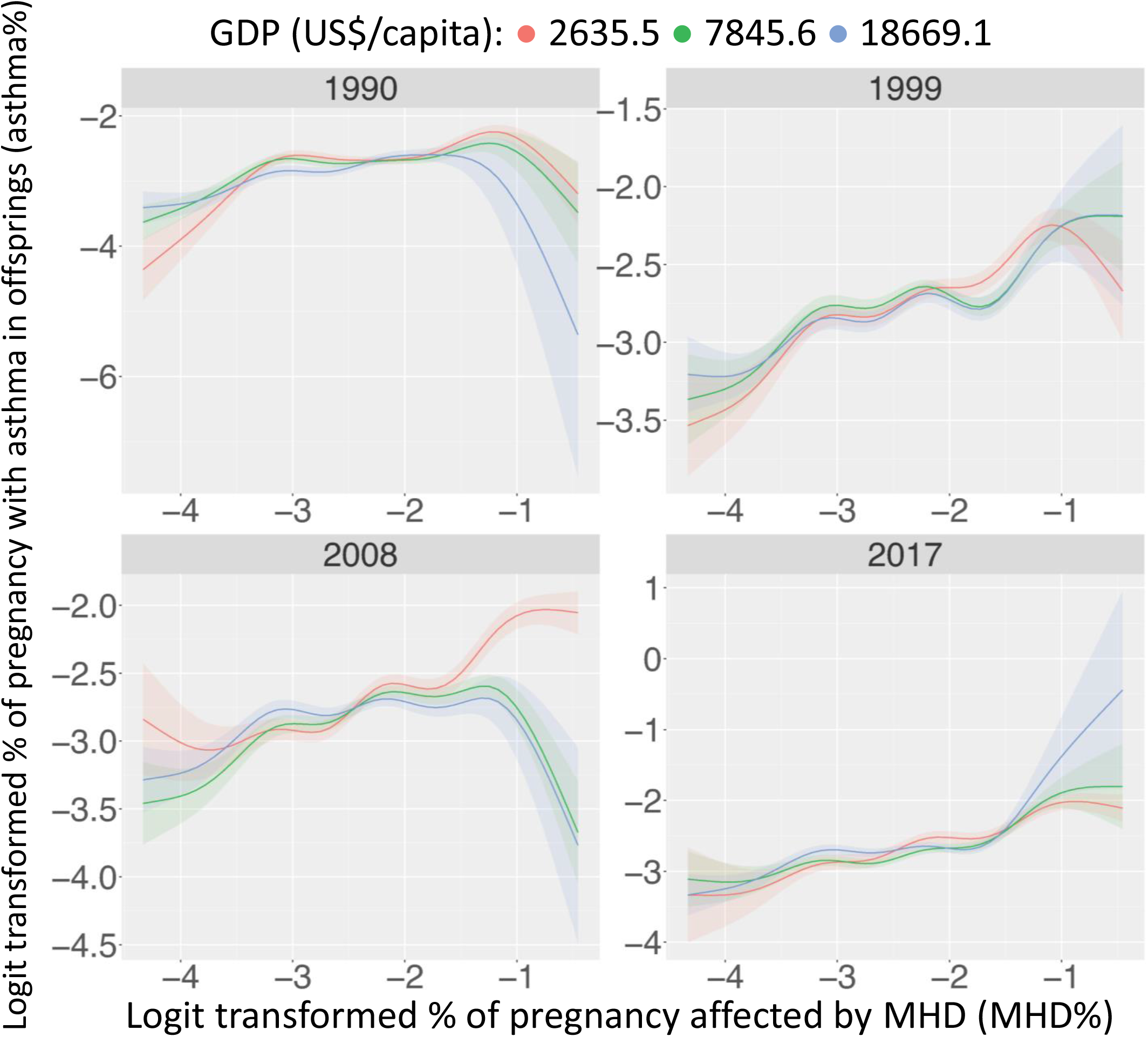
Predicted effects of logit transformed MHD%, GDP and time on the logit transformed asthma%. (See Supplementary Information for statistics)

Interestingly, aforementioned associations seemed to dissipate with age. For example, AD% for 10-14 years old exhibited less striking correlation with MHD% 12 years prior (Figure S1).

Collectively, we for the first time demonstrated positive associations between MHD% and offspring AD% and asthma% on a global scale.

Previous research primarily concentrated on preeclampsia, a severe form of MHD, and were typically in small observation cohorts. Our global analyses offered a more comprehensive overview while reducing potential biases like environments. However, global data for other allergic diseases beyond AD and asthma are lacking, which would be instrumental in expanding our findings. Further, stratified analyses for MHD and allergic disease severities could also yield deeper clinical insights.

The strongest association was found for 1-4 years AD relative to older ages and to asthma, latter being diagnosed usually after 2 years old. Hence, AD, the first condition within the atopic march^5^, is likely to best reflect the MHD influences. These impacts might be masked later in life by confounders, including nutrition and pollution. More comprehensive studies are thus warranted to inform allergic disease prevention.

Mechanisms underlying our findings remain unclear. We previously reported that preeclampsia impaired fetal regulatory T cell development^6^, crucial for atopy prevention, potentially explaining our observations. Investigating whether these mechanisms extend to other forms of MHD, and other immunopathology is necessary.

Overall, our study supports the Developmental Origins of Health and Disease (DOHAD) concept and showcases a novel methodology for DOHAD-related research, unravelling the maternal impacts on non-communicable diseases in offsprings.

## Data Availability

All data produced in the present study are available upon reasonable request to the authors

## Author contributions

*Concept and design:* Duan Ni, and Ralph Nanan

*Acquisition, analysis and interpretation of data:* Duan Ni, and Ralph Nanan

*Drafting of the manuscript:* Duan Ni, and Ralph Nanan

*Critical revision of the manuscript for important intellectual content:* All authors

## Acknowledgements

This project is supported by the Norman Ernest Bequest Fund.

