## Supplementary Information for "Global associations of maternal hypertensive disorders and offspring allergic disease burden"

### Supplementary Methods

*Data collection and processing*

Asthma, atopic dermatitis (AD), and maternal hypertensive disorder (MHD) disease burden data was downloaded from Global Burden of Disease (GBD) Study 2019 (https://vizhub.healthdata.org/gbd-results/). Crude birth rate (birth per 1000 people) and population data was from the World Bank databank (https://databank.worldbank.org/source/gender-statistics#). The gross domestic product (GDP) per capita data was obtained from the Maddison project[1]. GBD reported data from 1990 to 2019 and analyses were therefore based on this period of time. After collating disease and GDP data and excluding countries or timepoints with no data record, final data spanning 1990-2019 across about 150 countries were generated for analysis.

Pregnancy data per year was calculated based on crude birth rate and population data of each country. Percentages of pregnancies affected by MHD (MHD%) were calculated as the ratios of MHD cases per year and the pregnancies per year of the same country. As the peaks of incidences of asthma and AD are about 1-4 years old[2], this age group was focused for our analysis. Ratios of 1-4 years old asthma and AD cases per year versus the pregnancies 2 years prior of the same country were calculated as proxies of the percentages of pregnancies with offsprings developing asthma (asthma%) and AD (AD%) at 2 years of age. AD% and asthma% was compared with MHD% at the 2 years prior timepoint to decipher the link between MHD and offspring AD and asthma. Percentage data was all logit transformed before analysis.

*Generalized additive models (GAMs)*

Generalized additive models (GAMs)[3-5] feature in their capacity to interrogate multiple parameters simultaneously, as well as their potential interactions and non-linear effects, through the introduction of nonparametric smoothed functions. These functions are usually in forms of splines and provide flexible manners to evaluate non-linear associations.

Here, GAMs were run for AD% or asthma% using MHD% 2 years prior, GDP 2 years prior and time as predictors with the *mgcv* package and the "*gam*" function within[5,6]. In all models, countries from which the data originated were considered as random effects.

GAMs with aforementioned predictors and their different combinations, as well as a null model in which only the random effects of countries were considered were run. Using *mgcv* package[5,6], individual parameters were modelled as smooth terms using the "s()" function, while their interactions were modelled utilizing the "ti()" or "te()" function, given their different scales, like MHD% and GDP.

Modelling results were compared according to their Akaike information criterion (AIC)[7] and the one with the lowest AIC was selected.

Figure S1

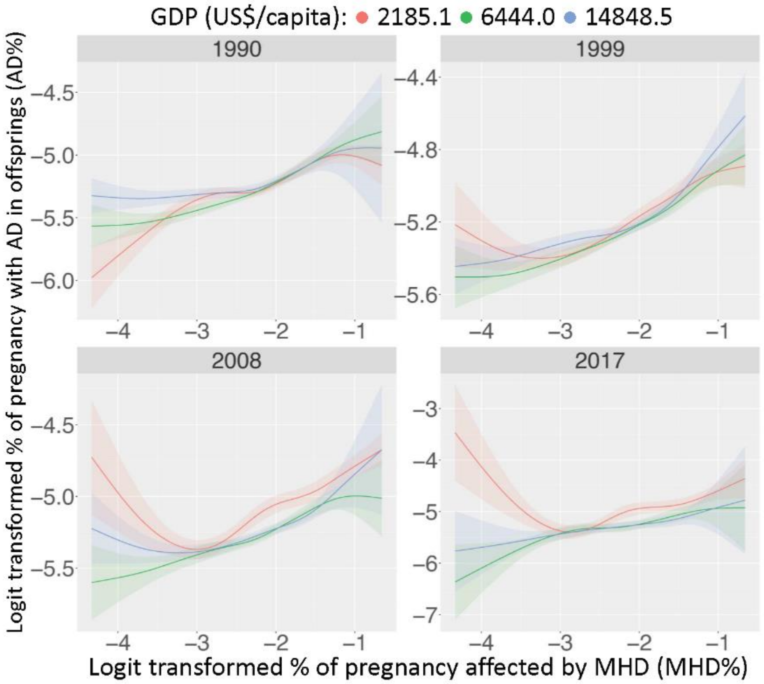


**Figure S1.** Predicted effects of logit transformed MHD%, GDP and time on the logit transformed AD% in 10-14 years old age group.

**Supplementary Tables**

Table S1-4 are GAM estimates and statistical outputs. The model estimates and related standard errors (Std.Error) were shown for parametric terms. For non-parametric smooth terms, their estimated and reference degrees of freedom (i.e. edf, sumEDF, Ref.df) and their test statistics were shown.

**Table S1.** Relative fit of GAM analyzing the predictors for AD%. AIC = Akaike information criterion. sumEDF indicates the degrees of freedom of the models. Model 16 was selected as the best model because of the lowest AIC. (related to Figure 1)

| GAM | AIC | sumEDF | Formula |
|---|---|---|---|
| 0 | -11055.96 | 157.91 | gam(AD% ~ 1 + s(Country, bs = "re")) |
| 1 | -11292.71 | 166.47 | gam(AD% ~ s(Country, bs = "re") + s(GDP)) |
| 2 | -11144.48 | 162.09 | gam(AD% ~ s(Country, bs = "re") + s(Year)) |
| 3 | -11545.51 | 169.48 | gam(AD% ~ s(Year) + s(GDP) + s(Country, bs = "re")) |
| 4 | -11579.22 | 176.77 | gam(AD% ~ te(Year, GDP) + s(Country, bs = "re")) |
| 5 | -12320.01 | 166.78 | gam(AD% ~ s(MHD%) + s(Country, bs = "re")) |
| 6 | -12372.69 | 171.04 | gam(AD% ~ s(MHD%) + s(Year) + s(Country, bs = "re")) |
| 7 | -12645.50 | 175.48 | gam(AD% ~ s(MHD%) + s(GDP) + s(Country, bs = "re")) |
| 8 | -12905.60 | 178.51 | gam(AD% ~ s(MHD%) + s(Year) + s(GDP) + s(Country, bs = "re")) |
| 9 | -12966.20 | 187.01 | gam(AD% ~ s(MHD%) + te(Year, GDP) + s(Country, bs = "re")) |
| 10 | -13094.57 | 182.37 | gam(AD% ~ te(MHD%, GDP) + s(Year) + s(Country, bs = "re")) |
| 11 | -12924.92 | 187.87 | gam(AD% ~ te(MHD%, Year) + s(GDP) + s(Country, bs = "re")) |
| 12 | -13523.58 | 263.55 | gam(AD% ~ te(MHD%, Year, GDP) + s(Country, bs = "re")) |
| 13 | -12523.69 | 178.17 | gam(AD% ~ te(MHD%, Year) + s(Country, bs = "re")) |
| 14 | -12883.89 | 180.03 | gam(AD% ~ te(MHD%, GDP) + s(Country, bs = "re")) |
| 15 | -13395.86 | 241.13 | gam(AD% ~ s(MHD%) + s(Year) + s(GDP) + ti(MHD%, Year, GDP) + s(Country, bs = "re"), data = complete |
| **16** | **-13674.22** | **274.60** | **gam(AD% ~ s(MHD%) + s(Year) + s(GDP) + ti(MHD%, Year, GDP) + ti(MHD%, Year) + ti(MHD%, GDP) + ti(GDP, Year) + s(Country, bs = "re"))** |
| 17 | -13352.09 | 217.49 | gam(AD% ~ s(MHD%) + s(Year) + s(GDP) + ti(MHD%, Year) + ti(MHD%, GDP) + ti(GDP, Year) + s(Country, bs = "re")) |
| 18 | -1359260 | 267.85 | gam(AD% ~ s(MHD%) + s(Year) + s(GDP) + ti(MHD%, Year, GDP) + ti(MHD%, GDP) + ti(GDP, Year) + s(Country, bs = "re")) |
| 19 | -13496.98 | 255.39 | gam(AD% ~ s(MHD%) + s(Year) + s(GDP) + ti(MHD%, Year, GDP) + ti(MHD%, Year) + ti(GDP, Year) + s(Country, bs = "re")) |
| 20 | -13667.62 | 258.97 | gam(AD% ~ s(MHD%) + s(Year) + s(GDP) + ti(MHD%, Year, GDP) + ti(MHD%, Year) + ti(MHD%, GDP) + s(Country, bs = "re")) |

**Table S2.** Estimated effects of MHD%, GDP per capita and year on AD%. (related to Figure 1)

| **Parametric coefficients** | | | | |
|---|---|---|---|---|
| | **Estimate** | **Std.Error** | **t value** | **Pr(>\|t\|)** |
| **(Intercept)** | -2.9400 | 0.1561 | -18.83 | <2e-16 |
| **Approximate significance of smooth terms** | | | | |
| | **edf** | **Ref.df** | **F** | **P value** |
| **s(MHD%)** | 8.741 | 8.981 | 147.113 | <2e-16 |
| **s(Year)** | 3.753 | 4.757 | 21.124 | <2e-16 |
| **s(GDP)** | 8.711 | 8.978 | 56.466 | <2e-16 |
| **ti(MHD%, Year, GDP)** | 61.936 | 63.442 | 9.609 | <2e-16 |
| **s(Country)** | 156.991 | 158.000 | 1590.785 | <2e-16 |
| R-sq.(adj) = 0.994 Deviance explained = 99.4% | | | | |
| GCV = 0.0025635 Scale est. = 0.002149 n = 4278 | | | | |

**Table S3.** Relative fit of GAM analyzing the predictors for asthma%. AIC = Akaike information criterion. sumEDF indicates the degrees of freedom of the models. Model 16 was selected as the best model because of the lowest AIC. (related to Figure 2)

| **GAM** | **AIC** | **sumEDF** | **Formula** |
|---|---|---|---|
| 0 | -6005.396 | 163.74 | gam(Asthma% ~ 1 + s(Country, bs = "re")) |
| 1 | -6046.657 | 167.44 | gam(Asthma% ~ s(Country, bs = "re") + s(GDP)) |
| 2 | -6084.177 | 168.63 | gam(Asthma% ~ s(Country, bs = "re") + s(Year)) |
| 3 | -6182.928 | 177.04 | gam(Asthma% ~ s(Year) + s(GDP) + s(Country, bs = "re")) |
| 4 | -6221.603 | 185.02 | gam(Asthma% ~ te(Year, GDP) + s(Country, bs = "re")) |
| 5 | -6372.593 | 172.60 | gam(Asthma% ~ s(MHD%) + s(Country, bs = "re")) |
| 6 | -6498.153 | 177.68 | gam(Asthma% ~ s(MHD%) + s(Year) + s(Country, bs = "re")) |
| 7 | -6466.578 | 181.16 | gam(Asthma% ~ s(MHD%) + s(GDP) + s(Country, bs = "re")) |
| 8 | -6629.696 | 186.90 | gam(Asthma% ~ s(MHD%) + s(Year) + s(GDP) + s(Country, bs = "re")) |

| | | | |
|---|---|---|---|
| 9 | -6683.406 | 194.13 | gam(Asthma% ~ s(MHD%) + te(Year, GDP) + s(Country, bs = "re")) |
| 10 | -6600.789 | 192.65 | gam(Asthma% ~ te(MHD%, GDP) + s(Year) + s(Country, bs = "re")) |
| 11 | -6577.306 | 195.48 | gam(Asthma% ~ te(MHD%, Year) + s(GDP) + s(Country, bs = "re")) |
| 12 | -7170.909 | 279.89 | gam(Asthma% ~ te(MHD%, Year, GDP) + s(Country, bs = "re")) |
| 13 | -6472.943 | 187.27 | gam(Asthma% ~ te(MHD%, Year) + s(Country, bs = "re")) |
| 14 | -6414.504 | 187 | gam(Asthma% ~ te(MHD%, GDP) + s(Country, bs = "re")) |
| 15 | -7067.771 | 240.75 | gam(Asthma% ~ s(MHD%) + s(Year) + s(GDP) + ti(MHD%, Year, GDP) + s(Country, bs = "re"), data = complete |
| **16** | **-7392.585** | **283.31** | **gam(Asthma% ~ s(MHD%) + s(Year) + s(GDP) + ti(MHD%, Year, GDP) + ti(MHD%, Year) + ti(MHD%, GDP) + ti(GDP, Year) + s(Country, bs = "re"))** |
| 17 | -6882.094 | 225.35 | gam(Asthma% ~ s(MHD%) + s(Year) + s(GDP) + ti(MHD%, Year) + ti(MHD%, GDP) + ti(GDP, Year) + s(Country, bs = "re")) |
| 18 | -7338.786 | 276.73 | gam(Asthma% ~ s(MHD%) + s(Year) + s(GDP) + ti(MHD%, Year, GDP) + ti(MHD%, GDP) + ti(GDP, Year) + s(Country, bs = "re")) |
| 19 | -7288.454 | 272.75 | gam(Asthma% ~ s(MHD%) + s(Year) + s(GDP) + ti(MHD%, Year, GDP) + ti(MHD%, Year) + ti(GDP, Year) + s(Country, bs = "re")) |
| 20 | -7300.799 | 270.62 | gam(Asthma% ~ s(MHD%) + s(Year) + s(GDP) + ti(MHD%, Year, GDP) + ti(MHD%, Year) + ti(MHD%, GDP) + s(Country, bs = "re")) |

**Table S4.** Estimated effects of MHD%, GDP per capita and year on asthma%. (related to Figure 2)

| **Parametric coefficients** | | | | |
|---|---|---|---|---|
| | **Estimate** | **Std.Error** | **t value** | **Pr(>\|t\|)** |
| **(Intercept)** | -2.5803 | 0.1222 | -21.11 | <2e-16 |
| **Approximate significance of smooth terms** | | | | |
| | **edf** | **Ref.df** | **F** | **P value** |
| **s(MHD%)** | 8.877 | 8.996 | 48.591 | <2e-16 |
| **s(Year)** | 4.665 | 5.809 | 24.885 | <2e-16 |
| **s(GDP)** | 8.836 | 8.991 | 17.044 | <2e-16 |
| **ti(MHD%, Year, GDP)** | 54.425 | 56.943 | 9.336 | <2e-16 |
| **s(Country)** | 162.943 | 163.000 | 682.351 | <2e-16 |
| R-sq.(adj) = 0.967 Deviance explained = 96.9% | | | | |
| GCV = 0.012593 Scale est. = 0.011933 n = 4592 | | | | |
